# Delphi: Deep Learning for Polygenic Risk Prediction

**DOI:** 10.1101/2024.04.19.24306079

**Authors:** Costa Georgantas, Zoltán Kutalik, Jonas Richiardi

## Abstract

Polygenic scores (PGS) are relative measures of an individual’s genetic propensity to a particular trait or disease. Most PGS methods use a regression framework for polygenic modeling and assume that mutation effect estimates are constant across individuals. While these assumptions simplify computation, they increase error, and PGS are particularly less predictive for under-represented genetic ancestries. We developed and provide Delphi (deep learning for phenotype inference), an individual-level deep-learning method that relaxes these assumptions to produce more predictive PGS. Delphi can integrate up to hundreds of thousands of SNPs as input and model non-linear SNP-SNP and SNP-covariate interactions. We compare our results with linear PGS models and a gradient-boosted trees-based method. We show that deep learning can be an effective approach to genetic risk prediction. We report substantial performance gains for a broad range of continuous phenotypes compared to the state-of-the-art. Furthermore, we show that Delphi tends to increase the weight of high-effect mutations. This work demonstrates an effective deep learning method for modeling genetic risk that also generalizes well when evaluated on individuals from non-European ancestries.

## Introduction

The total genetic component of common traits and diseases is attributable, at least in part, to a cumulative effect of a large number of mutations across the entire genome (1). Genome-wide association studies (GWAS) can identify univariate relationships between common single-nucleotide polymorphisms (SNPs) and a given trait. A GWAS output comprises an estimated effect size coupled with a P-value of association for each tested SNP. A single scalar indicating relative genetic risk can be obtained by summing up the number of alleles weighted by the estimated effect size of SNPs, with or without non-genetic risk factors (2). These so-called polygenic risk scores (PGS) are commonly used to quantify an individual’s genetic propensity for a particular trait or disease and have potential clinical applications in prevention, diagnosis, and treatment (3; 4; 5).

Methods for PGS estimation have evolved considerably in the past decade. It was first observed that including mutations below the GWAS statistical threshold would increase predictive power (6; 7). Taking linkage disequilibrium (LD) into account by either clumping and thresholding (C+T) or by using a shrinkage method also improved performance (8; 9). More recent work has included advances in statistical learning and an improved understanding of biology to increase the predictive performance of PGS. For instance, Bayesian approaches can also consider minor allele frequency (MAF) (10) or incorporate functional priors (11) to modify the effect size estimates. These methods generally yield only marginal improvements over one another and share similar limitations: effect estimates are unaffected by covariates and are linear in allele count.

PGS typically become significantly less predictive when applied to other less represented ancestries (12). This performance drop can be partly attributed to differences in allele frequencies between cohorts and to other genetic and environmental factors (13). These limitations hinder the application of PGS in medical settings (14), and this performance gap can only be bridged with additional data collection from underrepresented ancestries. Multiple approaches have been proposed to increase the generalizability of PGS, for instance, by aggregating results from multiple GWAS studies (15; 16) or prioritizing functional variants (17). Recently, increased prediction performance was observed through the use of a gradient-boosted model taking a standard PGS and a selection of high-impact SNPs as inputs (18). Very recently, GenoBoost (19), another gradient-boosted approach, showed improved performance by modeling non-additive mutation effects.

Deep learning (DL) enables learning complex patterns directly from large labeled datasets with minimal assumptions. In genetics, DL has been applied to many problems, such as variant calling (20), motif discovery (21), and image-derived phenotyping for GWAS (22; 23). Explainable DL approaches (24) could provide additional insight into the genetic factors influencing the disease. Abe et al. recently constructed a knowledge graph (25) to generate text-based explanations for individual variants. Using deep learning for genetic risk prediction could provide unique advantages, as overparameterization has recently been shown to improve generalization (26), which is important for PGS to be applicable in underrepresented populations. Deep learning offers an advantage over conventional PGS methods because it does not depend on predefined assumptions regarding effect estimates. Using deep learning on genetic data enables evaluation of the potential gains associated with relaxing these assumptions.

Using DL for PGS estimation has been attempted before, although a lot of proposed approaches consisted in using shallow networks (max. 4 fully connected or convolutional layers) on a small set of SNPs (max. 10K) (27; 28; 29; 30; 31; 32). In those examples, DL was shown only marginally to improve results, if at all. For instance, Badré et al. (30) found that including 5273 high-impact SNPs in a deep neural network slightly improved the predictive performance of PGS for breast cancer over logistic regression, and including more SNPs did not improve performance. Zhou et al.(32) showed that a small neural network with three fully connected layers improved Alzheimer’s disease genetic risk prediction in a small (N ≈ 10K) cohort. Direct comparison between methods is challenging because no open benchmark datasets are currently available. As the explained variance depends on phenotypic variance, differences in predictive performance may arise from differences in datasets. Differences in genotype panels can also significantly affect predictive performance.

In this work, we propose Delphi (deep learning for phenotype inference), a deep learning method that alleviates some of the issues of PGS mentioned above by tuning risk score estimates in a data-driven and hypothesis-free manner. In contrast to previous methods, we use a transformer architecture to capture non-linear interactions. Unlike other approaches, effect estimates are modified during inference before the summation, allowing them to depend on covariates such as age and sex, and other mutations. Our method can fine-tune effects from any classical PGS method such as LDpred2 (8) and Lassosum (9). We compare our method to other linear PGS models and an XGBoost-based benchmark that allows for non-linear interactions. Finally, we assess the accuracy of our predictions for our method on individuals from multiple ancestries.

## Results

### The Delphi Method

At a high level, Delphi (Figure 1) uses genotyping and covariate information to learn individual-specific perturbations of mutation effect estimates. Our approach contains two main steps. (1) The dataset is split into training, validation, and test sets before pre-processing. PGS effect sizes are estimated using standard PGS techniques, and genotyping data are converted into a format that enables fast loading during training. (2) In the training step, a covariate model based on gradient-boosted trees (33) estimates the phenotype from age, sex, and genetic principal components, and a deep neural network learns to perturb individual effect sizes for all mutations included in the PGS summation according to Equation 1. The modified effect sizes are then summed to form a personalized PGS in which covariates and the presence of other mutations are accounted for in the personalized effect estimates. The covariate model outputs and the PGS summation are finally linearly combined to form the final prediction.

**Figure 1.**
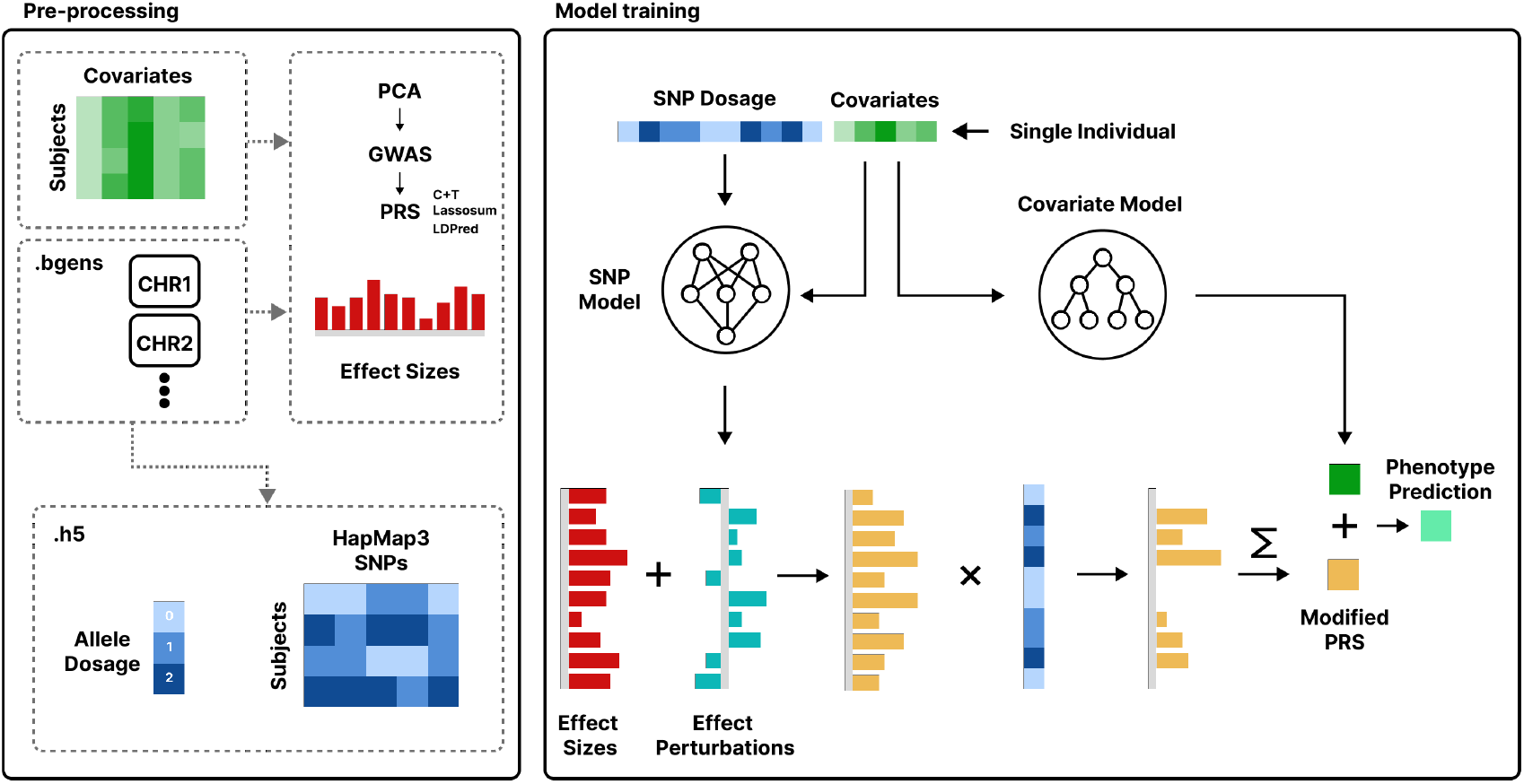
Overview of the Delphi method. The dataset is split into training, validation, and test sets before pre-processing. A GWAS is conducted on the training set for the phenotype of interest, followed by a PGS method. A transformer neural network learns to modify the individual-level effect size estimates during training depending on other SNP allele counts and covariates in the training dataset. Model selection for the neural network and the PGS methods is done using the validation set. Modified effect sizes are aggregated with the predictions of a boosted trees covariate model to form a new PGS. All prediction results are evaluated on the held-out test set.

We found that training the network directly on the phenotype led to unstable and divergent optimization, due to the high dimensionality of the genetic input space, the small marginal contribution of individual SNPs, and the fact that phenotypes are not fully determined by the observed inputs. We further investigate the sources of this training difficulty using synthetic data; see Section Synthetic Experiments. To address this issue, we use baseline effect size estimates ***β*** = {*β*_*j*_} obtained from a classical PGS method to guide the neural network’s predictions.

Specifically, the output of the transformer is decoded through a linear layer to match the dimensionality of the baseline effect sizes, and the network predicts individual-specific perturbations to each effect estimate using a tanh activation function:

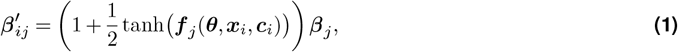

where ***β*** denotes the baseline effect estimates from the PGS, 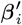 are the individual-specific modified effects, ***f*** (·) is the neural network parameterized by ***θ***, and ***x***_*i*_ and ***c***_*i*_ represent the genotype features and covariates for individual *i*, respectively.

Each modified effect 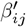 represents an individual-specific adjustment of the baseline SNP effect that depends on both covariates and the configuration of other SNPs. In this formulation, SNP–SNP and SNP–covariate interactions are implicitly captured by allowing effect sizes to be functions of the full input (***x***_*i*_, ***c***_*i*_). The individualized effect estimates are then summed to form the modified PGS prediction of the neural network,

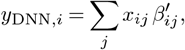

Importantly, the network output ***f*** (·) has the same dimensionality as ***β***, and the modulation of effect sizes is performed via element-wise multiplication. By construction, if the network outputs zero for all SNPs, the resulting score reduces exactly to the unmodified PGS. We found that using baseline effect estimates as a guide during training is essential for obtaining stable and predictive neural network models.

### Synthetic Experiments

We observed that standard neural network architectures had issues producing accurate predictions when including a large number (>10K) of SNPs as input, which was also observed in other works (28). We believe the challenge arises from the extremely small, largely additive per-SNP effects, which make it difficult for the neural network to extract a meaningful signal when trained directly on all variants. To study this pattern more deeply, we conducted two sets of synthetic experiments; the first, to highlight cases where the Delphi architecture can be advantageous in lieu of direct phenotype prediction, and the second to show that performance improvements can be obtained on realistic synthetic phenotypes.

Our first set of synthetic experiments compared the behavior of neural network in a synthetic dataset with a large number of features and an outcome that is mostly additive in nature. The phenotype is defined as follows:

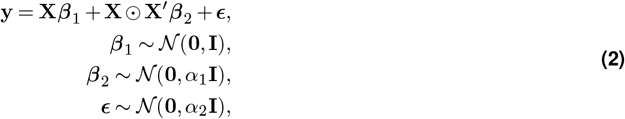

where **y** ∈ ℝ^*N*^ is the synthetic phenotype for *N* subjects, **X** ∈ {0, 1, 2}^*N* ×*M*^ is a matrix of randomly drawn integers between 0 and 2, with *N* subjects and *M* variants. ***β***_1_ ∈ ℝ^*M*^ is the vector of effect sizes for the variants. **X**^′^ is a random column-wise permutation of this matrix. **X** ⊙ **X** is an element-wise multiplication (Hadamard product) to simulate linear interactions, with associated effect sizes ***β***_2_ ∈ ℝ^*M*^. The identity matrix **I** ∈ ℝ ^*M* ×*M*^ is an isotropic covariance matrix with variance 1. Additionally, we set a fixed causal fraction of inputs with effect *β*_*i*_ = 0. The variance of interaction effect sizes (*α*_1_) and the variance of the error (*α*_2_) are tunable scalar parameters.

We compared direct prediction, as is usually done in deep learning, to first estimating the average effect of features with individual linear regressions on the training set, as is done in GWAS, and then learning variations of effect estimates that depend on the input to capture non-linear interactions, as described in Equation 1. We present a fully reproducible minimal example that compares the standard fully connected, convolutional, and transformer architectures on a synthetic phenotype with additive and non-additive components.

We trained three neural network architectures to predict phenotype from variants, to evaluate the effectiveness of the Delphi approach compared to direct training on variants: a fully connected network, a convolutional network, and a transformer. Delphi effect estimates 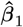 were obtained by linear regression, obtained exclusively on training data, to simulate GWAS results. All architectures were trained for 100 epochs with a learning rate of 0.001. We varied *α*_1_ between 0 and 5 and set *α*_2_ = 0.1 for all experiments. Delphi neural network architectures were only modified at the last layer to match the input sizes.

Results are shown in Supplementary Figure 11, we consistently observed better performance for all tested architectures with a dataset of size *N* = 10^′^000 and a causal fraction of 0.5 and *M* = 10^′^000 features. We also reproduced the same experiment with a smaller causal fraction of 0.1, in which case we observed that Delphi was better than the baseline when the interaction factor was sufficiently small, i.e *α*_1_ *<* 2 (Supplementary Figure 11). In addition, our proposed method converges much more rapidly, requiring less than 10 epochs of training versus 100 or more for other methods. Our approach is specifically tailored to objectives that are primarily additive in nature and appears to be beneficial in this setting when the number of features is sufficiently large. We also constructed a monogenic synthetic phenotype with a single predictive input. In this situation, Delphi performed much worse than benchmarked neural networks, as shown in Supplementary section 8.

In our second set of synthetic experiments, we used HAPNEST (34), a library for efficiently generating individual-level genotypic and phenotypic data. Genotypes composed of 1,329,052 HapMap3 SNPs were simulated for 100,000 individuals of European ancestry. HAPNEST effect sizes are drawn from a Gaussian distribution with mean 0 and variance depending on heritability, MAF, local linkage, and functional annotations. We constructed three phenotypes in which genetic effects accounted for 50% of the phenotypic variance. The first two, P1 and P2, were obtained directly from the HAPNEST simulation with polygenicity set to 0.05 and 0.005, respectively. For each individual, the third phenotype was constructed to introduce widespread SNP by covariate interaction effects, by combining the first two phenotypes in the following manner:

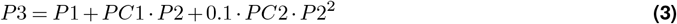

Principal components (PCs) and phenotypes are Z-normalized on the training set before constructing the new phenotype. As the polygenicity of the first phenotype is greater than that of the second, this effectively adds a perturbation effect to a portion of the SNPs. Once the data were generated, we replicated the experiments we conducted on the UK Biobank (see Methods), testing the behavior on three independent train/validation/test splits (70%, 10%, 20%). The first 10 PCs were used as covariates for Delphi and for the linear regressions. The GWAS were done by combining training and validation sets. More details on the generation of synthetic phenotypes and genotypes with HAPNEST can be found in Supplementary Section 8.

Predictive performance was compared across three methods, C+T, LDpred2, and our proposed approach. Results are summarized in Figure 2. The relative difference in explained variance between Delphi and the next-best method is 14.2%, 6.0%, 25.7% for the three phenotypes. For the two purely additive phenotypes, performance was comparable across methods, with no substantial differences observed. In contrast, for the interaction-driven phenotype, the proposed approach showed a clear improvement over C+T and LDpred2. This gain was consistent across runs and underscores the Delphi architecture’s ability to capture covariate-dependent genetic effects that are not explicitly modeled by standard polygenic risk score methods. These results indicate that while conventional approaches remain competitive for simple additive architectures, modeling strategies that directly incorporate interactions can provide substantial benefits in settings where such effects are widespread.

**Figure 2.**
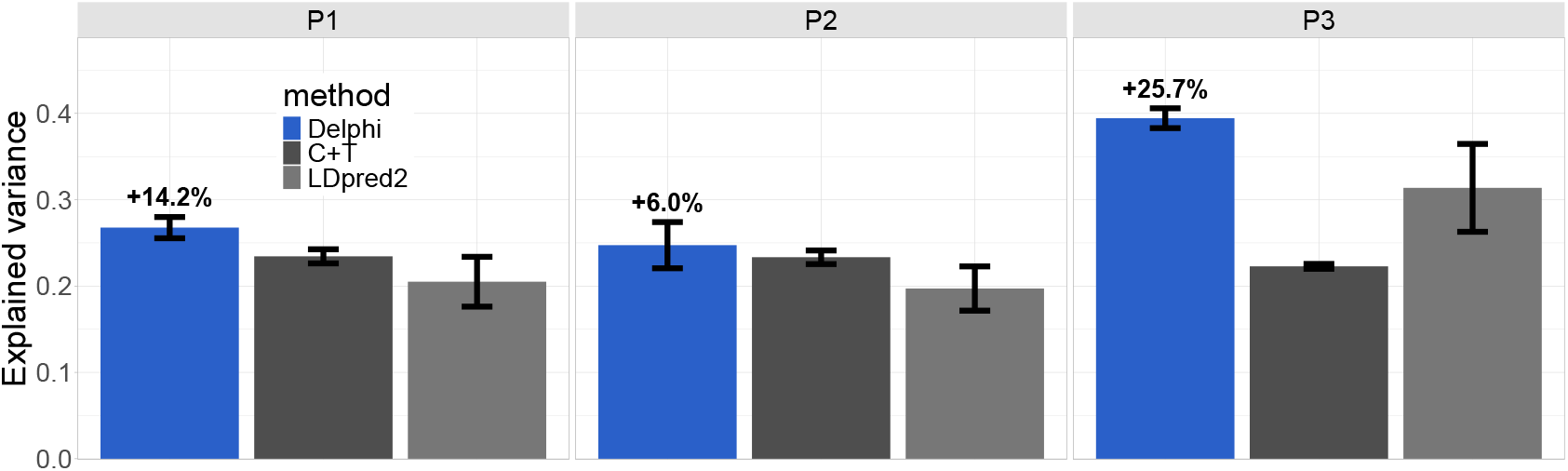
Comparison of prediction performance between Delphi, clumping and thresholding (C+T), and LDpred2 on three synthetic phenotypes generated with HAPNEST. Results are averaged on three independent splits (the same for all methods), P3 contains an additional PC dependent interaction effect on a portion of SNPs.

### GWAS and PGS

407’015 unrelated UKBB subjects were randomly split into different sets. The training set (325’606 subjects) was used for principal component analysis (PCA), GWAS, PGS computation, and deep neural network training. The validation set (5000 subjects) was used for PGS validation and model selection after training. The held-out test set (76’409 subjects) remained unseen until the final evaluation. We only considered the 1,054,330 SNPs from the HapMap3 set (35) with an INFO score *>* 0.8 and MAF *>* 0.01. PCA on the genotype matrix was used to capture population structure.

GWAS for all phenotypes only included subjects within the training set from British-white ancestry (UKBB field 22006) to reduce spurious associations, and any subjects further than three standard deviations away from the first six principal components were removed. Sex, age, and the first 20 principal components were used as covariates. Classical PGS methods use LD, MAF, and other measures to reweight the effect estimates. We found some performance improvement by using these re-weighted effect estimates as a baseline instead of the GWAS summary statistics. PGS were obtained with three different methods: C+T, Lassosum (9), and LDpred2 (8). All steps of the preprocessing, including GWAS, PCA and PGS estimation were performed using the bigsnpr (36) R package (version 1.9.10).

### Learning perturbations of mutation effects

The second step consists of learning individualized effect perturbations. As in GWAS, covariates were age, sex, and the first 20 PC loading. Before transformer training, an XGBoost model is fitted on covariate data from the training set and is referred to as the covariate model. Separately, the genotype data was converted from.bgen files to a hierarchical format that allows for fast data retrieval of all HapMap3 SNPs of a small number of subjects. We trained the neural network on the residuals of this model, which made convergence easier when some covariates had a high impact on the phenotype. The neural network’s architecture was a standard 8-layer transformer with variable sequence length depending on the number of input SNPs. SNPs were aggregated into fixed-size groups and linearly mapped to form a sequence of embeddings of size 512. In addition, covariates were included as the first embedding in the sequence, and zero padding was used when necessary. The transformer’s output was then mapped back into a vector the size of the number of input SNPs. This vector represents individualized variations in the SNP effect. As in traditional PGS methods, these modified effects were then summed up and linearly mapped in combination with the output of the covariate model to form a final prediction. A graphical overview of the method is presented in Figure 1.

### Baseline PGS results

Three PGS methods (C+T, Lassosum2, and LDpred2) were compared to provide baselines. We compared the proportion of explained variance (EVR) for all phenotypes. Predictions were made with a linear or logistic model, using age, sex, and the first 20 genetic PCs as covariates and the estimated score. LDpred2 outperformed the other two tested methods on all tested phenotypes. Thus, we chose the effect estimates from LDpred2 as the baseline for our method in all further analyses. We also compared the performance of two variants of LDpred2: LDpred2-grid and LDpred2-auto. We found that LDpred2-auto was superior to LDpred2-grid for all tested phenotypes and therefore used this variant. Performance for the tested methods on ten continuous phenotypes is shown in Supplementary section 1.

### Trait Prediction

We evaluated the performance of Delphi on ten continuous phenotypes, with the same train/validation/test splits used for benchmarking other methods. Phenotypes were selected based on their appearance in other similar works (18), because they were known to be polygenic, and were differently impacted by covariate information. We use the explained variance as a performance metric, and provide results for other metrics (normalized MAE) in Supplementary section 2. We also compared Delphi with linear and Lasso regressions and an approach using XGBoost to modify effect sizes (18) with the base weights from LDpred2. Hyperparameters (number of boosted trees and regularization parameters) for all three methods were tuned with three-way cross-validation on the same validation set. For the XGBoost model, multiple P-values for the inclusion of high-impact SNPs were tested, see section Model performance evaluation and comparison to existing methods. Results showed that Delphi yielded lower error than other approaches on all phenotypes. Figure 3 provides detailed results, and performance in terms of normalized MAE (w.r.t to the worst performing models) are show in Supplementary Figure 2. Performance tables are provided in Supplementary section 5. It should be noted that the explained variance from benchmarked PGS methods is lower than the ones shown on the validation set in Figure1, as the validation set was used for parameter tuning on this set for all three methods. We have also obtained similar performance improvements from Delphi using REGENIE PGS effect sizes instead of LDpred2; see supplementary section 7 for details.

**Figure 3.**
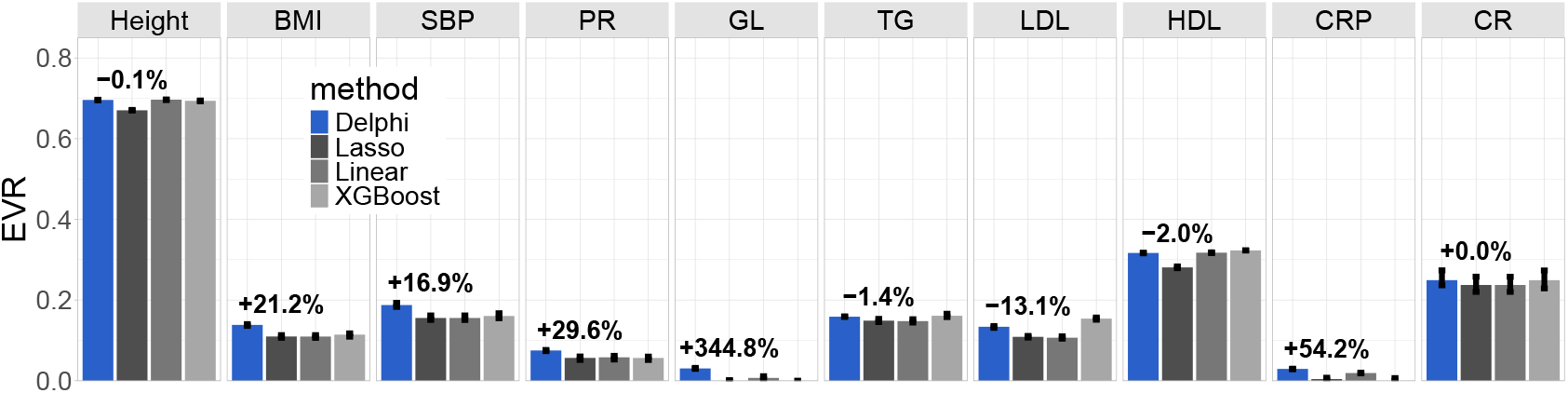
Accuracy of polygenic predictions for ten phenotypes in the UK Biobank (test set, 76,409 participants). We report results for a linear PGS model based on LDpred2 effect sizes with covariates (age, sex, top 20 PCs) included as predictors, lasso regression with the same covariates, an XGBoost model including the allele count of multiple high-impact SNPs as input, and our method. EVR: explained variance, BMI: body mass index, SBP: systolic blood pressure, PR: pulse rate, GL: glucose, TG: triglycerides, LDL: low-density lipoproteins, HDL: high-density lipoproteins, CRP: C-reactive protein, CR: creatinine.

We compared Delphi prediction to the next best method, XGboost, in terms of the distribution of errors. Predictions from Delphi showed, in general, lower absolute error than XGboost predictions (Supplementary section 2). Delphi generally tended to have fewer large prediction errors, as shown by the ratio of quartile difference between the two methods (Supplementary Figure 3).

The relative increase in the percentage variance explained compared to the benchmarked methods was 21.2% for body mass index, 16.9% for systolic blood pressure, and 29.6% for pulse rate. Delphi performs consistenlty better on phenotypes that are difficult to predict, such as glucose levels and CRP (2%, and 1% gain in absolute EVR, respectively). Performance on other phenotypes was similar, or lower in the case of LDL (-13.1% relative EVR). we also compared the predictive accuracy of different components that make up Delphi. We evaluated the predictive performance of the covariate model, the baseline unmodified effect estimates, and the effect estimates individually modified by Delphi. The baseline and modified effect estimates were summed and weighted as would be in a traditional PGS, and evaluated uncorrected for covariates. As shown in Figure 4, the Delphi-tuned effect estimates are consistently more predictive than the baseline effects. This means that the genetic part of model’s outputs are consistently more predictive after training.

**Figure 4.**
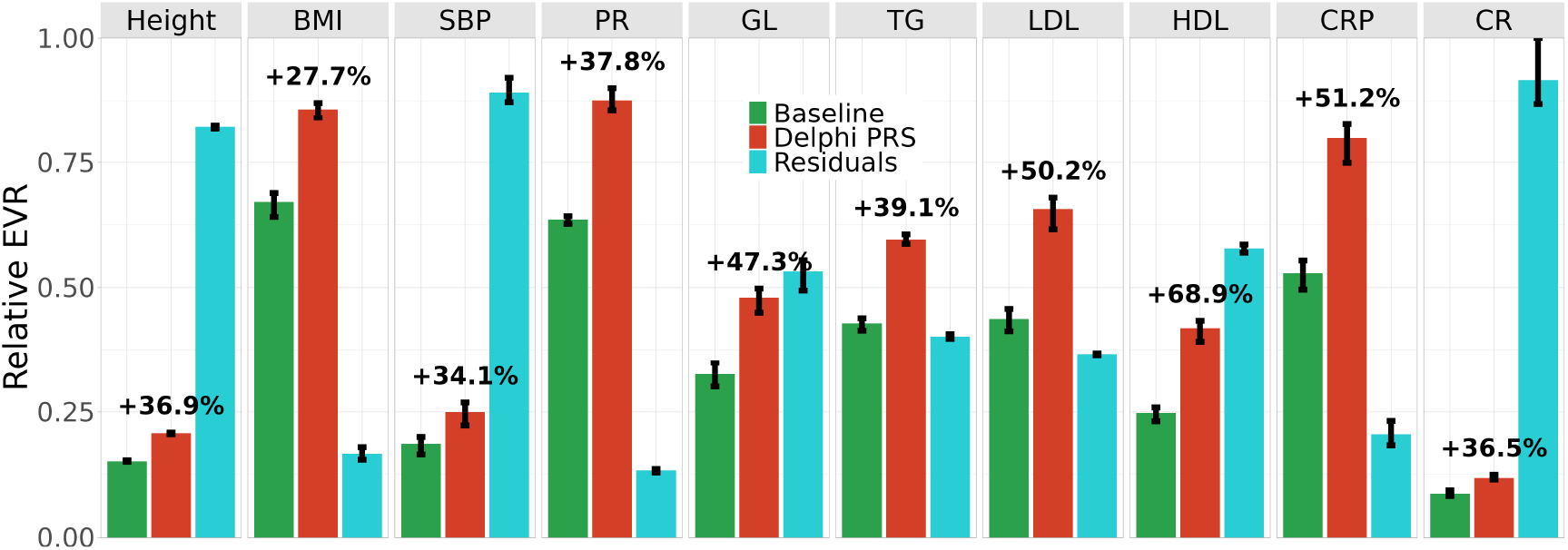
Relative explained variance w.r.t the combined prediction for different components of the Delphi model: the linear PGS based on LDpred2 effect sizes taken as input by Delphi (“baseline”, not including covariates), the SNP effects after modulation by Delphi (“Delphi PGS”), the variance explained by covariates not captured by the PGS component (“residuals”). Delphi’s effect modifications consistently explain more variance than the baseline PGS.

### Performance comparison on multiple ancestries

We compared the prediction performance difference on multiple subsets of the test set based on ancestry. Ancestries were defined according to previous work (37), and were based on PCs computed within the UK Biobank and individual information on the countries of birth. We show the results on the held-out test sets for Nigerian (N ≈ 770), Indian (N≈ 1260), and Chinese (N ≈ 350) ancestries in figure 5 and the others in Supplementary Figure 5. Despite the low number of subjects in each group, Delphi outperforms other tested approaches on most phenotypes for all recorded ancestries.

**Figure 5.**
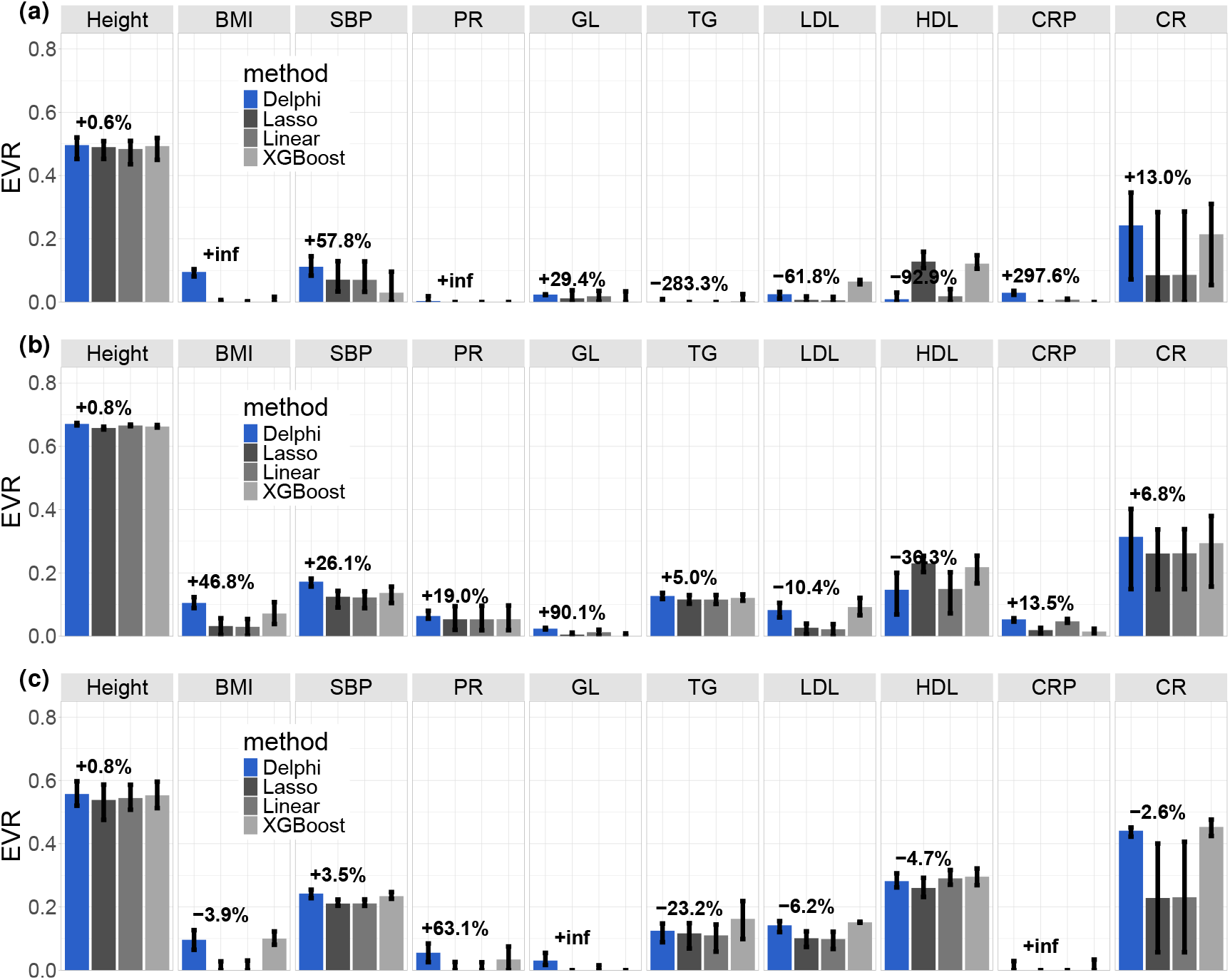
Accuracy of polygenic predictions for ten continuous phenotypes in the UK Biobank for three different ancestries on the test set. a) Nigerian, b) Indian, c) Chinese. Ancestries were defined according to previous work (37), which used populations of the 1000 Genomes Project as reference.

We also compared performance on the subset of the test sets with non-British white ancestries (N ≈ 13K, depending on split and phenotype) to assess the performance change due to ancestry on a large number of participants. The same exact splits were used for the benchmarks and for our approach. Ancestry was determined according to Field 22006, which indicates subjects who self-identified as ‘White British’ and have very similar genetic ancestry based on a principal components analysis of the genotypes. Individuals with non-British white ancestries accounted for approximately 13% of the dataset. Results are shown in Supplementary Figure 5.

The relative increase in the percentage variance explained compared to the tested benchmarks on the set with non-British white ancestries was 33.5% for body mass index, 30.2% for systolic blood pressure, and 66.6% for pulse rate. Performance drops were observed for LDL and HDL (-21.5%, -10.8% respectively). Notably, the EVR is higher for non-British white individuals for some phenotypes (BMI and LDL). This predictive gain is due to the reduction of the total variance and is not reflected in the mean absolute error (see Supplementary section 2). Results on the British white set were otherwise very similar to the total set shown in Figure 3, as these individuals represent approximately 95% of the held-out test set.

### Observed Trends in Effect modulation

We observed interesting patterns when inspecting the average effect modulations before the summation. As shown in Figure 6, Delphi tends to down-weight the absolute effect of SNPs with low absolute effect. Interestingly, we do not observe the same trend when grouping SNPs by minor allele frequency decile. Other SNP-heritability estimation methods such as LDAK (38) include MAF, LD estimates and functional anotations to refine predictions. As LDpred2 modifies the effect estimates before any modification by the deep neural network, we expect the LD structure to be included in the effect estimates.

**Figure 6.**
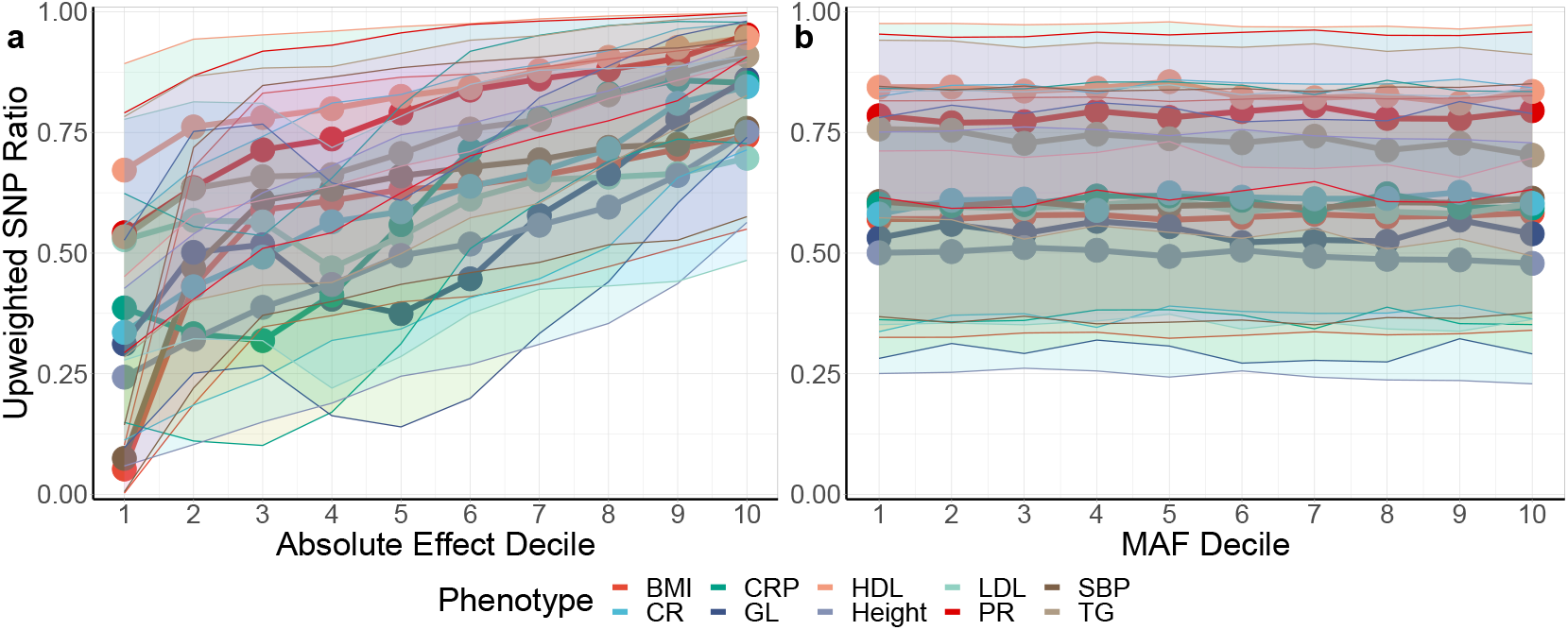
a) The ratio of up-weighted SNPs in Delphi by absolute effect size decile was estimated with LDpred2 for ten phenotypes. This ratio was computed by dividing the number of of SNPs with higher estimated effect on average on the test set with the total number of SNPs for search decile. b) Ratio of up-weighted SNPs in Delphi by minor allele frequency. Delphi tends to down-weight the absolute effect of SNPs with low absolute effect, while SNP effect is not generally modulated by MAF.

## Discussion

We introduced Delphi, a deep-learning-based method for trait prediction from genetic data. We demonstrated that deep learning enhances the predictive power of polygenic risk scores. Treating genetic risk estimation as a deep prediction problem allowed us to relax the usual assumptions of traditional PGS methods, yielding significant performance improvements on multiple phenotypes over previous PGS computation methods.

Several studies have tested deep learning approaches for phenotype inference from genetic data. These approaches all have a similar structure: use GWAS summary statistics to select a subset of SNPs, then use these as input for the neural network. Uppu et al. (27) used a 3-layer feed-forward network applied to breast cancer data. To our knowledge, this study contains the first use of a neural network for genetic risk prediction. In contrast, Bellot et al. (28) did not find any performance gain when comparing convolutional and fully connected neural networks to traditional methods. Recently, Huang et al. proposed DL-PRS (29), a method that also uses a shallow network to predict COPD, achieving no statistically significant performance gains over traditional methods on UKBiobank data. A similar approach has been used by Badre et al. (30) with a 4-layer FC neural network on breast cancer data. Very recently, Zhou et al. (32) used a graph neural network for Alzeihmer’s prediction by constructing a graph from a few correlated locis. Elgart et al. (18) showed the strongest evidence for the superiority of non-linear methods for PGS by using gradient-boosted trees. This publication obtained robust results across multiple traits, which motivated our study.

All previous works that used fully connected or convolutional layers observed a cap in performance gain when including a large (*>* 10K) number of SNPs as input. Some works mention memory usage as the main limitation for using a large number of input SNPs (30; 24; 39); we show through synthetic experiments and in the UK Biobank that computational cost is not the only problem with this approach. We observed that training becomes difficult as most SNPs have a minimal individual impact on the phenotype and lack exploitable patterns. This is particularly a problem for PGS, which can include hundreds of thousands of SNPs.

Other works use sparsely connected networks designed from known gene-SNP interactions (39; 24). Sparsely connected layers were used precisely because fully connected layers would have been computationally infeasible. Notably, Sigurdsson et al. (40) constructed a genome-local network (GLN) to model non-additive effects. While this approach may have other benefits, such as explainability, the constraints of the neural architecture imply that there can be only a limited number of pre-specified interactions between SNPs and that SNP-covariate interactions are not taken into account. In contrast, deep transformer networks have achieved better results across many other data modalities, including text, images, and tabular data. To guide the neural network towards meaningful predictions, we chose to perturb the estimated effect sizes rather than predicting the phenotype directly. As a result, we can effectively integrate hundreds of thousands of SNPs as input in a transformer-based neural network, which would not be feasible with direct prediction.

We have also shown that our method generalizes well when evaluated on individuals from non-European ancestries, although our training set is composed of 95% European. This is an essential point for the success of PGS in any clinical setting, or their application can potentially reinforce ancestry bias (41). Our approach could be combined with other methods for the standardization of PGS, for instance, by combining summary statistics from multiple GWAS studies (42) or through some debiasing measure (43). The performance and fairness of PGS is an ongoing problem and requires more data acquisition from non-European cohorts. To reduce these disparities, it is necessary to assess and maintain prediction performance for all populations thoroughly. Delphi is currently not designed as a cross-ancestry PGS method, but could potentially be extended by incorporating multiple sources of effect estimates. We leave the extension of this approach for further research.

Our study presents several limitations. The high dimensionality of the data, combined with the sizeable but still limited number of samples, necessitated trade-offs to maintain consistent prediction performance across all traits. Similar to other PGS methods, the most crucial hyperparameter to tune is the minimum threshold probability for SNP inclusion. This threshold also affects the number of SNPs we batch in a single embedding vector, and training can diverge when including too many non-significant SNPs. A threshold of *>* 1% in MAF also limits our ability to generalize to other ancestries, but is required for estimating effect sizes. Finally, our approach also takes significantly more computational power and time than the other compared methods, and results in less interpretable outputs due to the black-box nature of deep learning models.

We observed that the benefits of our method compared to benchmarks would depend on the phenotype. We suppose that Delphi works particularly well for phenotypes that are highly polygenic and known to exhibit SNP-sex and/or SNP-PC interactions, such as height (44). Delphi tends to increase the influence of SNPs with high effect estimates and downweights those with low effect estimates. Similar heuristics have been shown to improve heritability estimates by tuning effects based on minor allele frequency (MAF) (10). This observation might indicate that the absolute effect size may be an additional parameter of interest for future Bayesian methods. Although MAF is correlated with effect size, we found no such association by inspecting variations of modulation and MAF quantiles, as shown in Figure 1. Unfortunately, we also found that the effect estimates for individual SNPs varied substantially across data splits, making the interpretation of SNP effect modulation challenging, as the variations in the neural network were much smaller than the differences due to data randomization. This limitation might be alleviated by including summary statistics from another cohort.

## Methods

### UK Biobank

The UK Biobank (UKBB) (45) is a large-scale ongoing prospective study including over half a million individuals from across the United Kingdom. Participants were first recruited between 2006 and 2010 and underwent extensive testing, including blood biomarkers, health and lifestyle questionnaires, and genotyping. Longitudinal hospitalization data for any disease represented by an ICD-10 code is also provided between the recruitment date and the present time. UKBB contains genotypes for 488,377 individuals at the time of download (March 2023), 409,519 of which are from ‘white British’ ancestry. Ancestry was inferred using the data field 22006, which uses self-reports and principal component analysis of the genotypes. Variant quality control included the removal of SNPs with imputation info score *<* 0.8 and retaining SNPs with hard-call genotypes of *>* 0.9 probability and MAF *>* 0.01. To reduce the initial dimensionality of the data, we only considered 1,054,330 HapMap3 (HM3) (46) SNPs as they have shown to be a sufficient set for traditional PGS methods (47) and are the standard set for polygenic risk score evaluation.

### Data Splits and Phenotypes

We evaluated the performance of our method on all ten traits using three independent train/validation/test splits. For the quality control of our samples, we only considered 407,008 subjects used in the principal components analysis of the UKB dataset (field 22020). These subjects are unrelated, did not withdraw consent from the study, and passed some genotyping quality control tests. Subjects were not selected based on ancestry at this stage. We used 80% (325’640) of the dataset for training, 5000 subjects for validation, and the rest (76’409 subjects) for testing. Some individuals were further removed depending on missing data for each phenotype. We kept this exact split for the preprocessing (GWAS and PGS) and the training of the neural network. The same training/validation/test sets were used for the benchmarked methods. The validation set was used to select the best hyperparameters for polygenic risk scores and benchmark algorithms and to stop the deep neural network training. We assessed the performance of our method on ten continuous phenotypes. BMI, height, SBP, LDL, and C reactive protein values were taken directly from the UK Biobank first time point measurements.

The LD reference panel used for LDpred2 was previously computed (47) with some individuals from the test set. The choice of LD reference panel was shown to have a limited impact on performance (47), and the same weights from LDpred2 were used for all benchmarked methods. Finally, this panel did not contain individuals from non-British white ancestry, ensuring that performance results on non-British white (see section Performance comparison on non-white British for multiple phenotypes) are unbiased.

### PCA and GWAS

PCA eigenvectors were obtained from 1,054,330 HapMap3 SNPs using only genotype information from the training set for each data split. As recommended (48), we used a truncated PCA method with initial pruning that iteratively removes long-range LD regions. GWAS were computed on all HM3 SNPs and the training set was pruned by removing individuals with no British white ancestry (field 22006) and who were beyond two standard deviations of the Mahalanobis distance of the first 6 PCs. This additional subject selection was only applied for the GWAS to prevent spurious relationships that can arise with heterogeneous cohorts. Covariates for the regression included age, sex, the first 20 principal components, age^2^, age · sex and age^2^ ·sex. PCA and GWAS were computed using the bigsnpr (36) R package (version 1.9.10).

### PGS Computation

Polygenic risk scores were computed for each phenotype and data split using clumping and thresholding (C+T), lassosum2, and LDpred2. In C+T, correlated variants are first clumped together, leaving only the ones with the lowest P-values while others are removed. We used 50 P-value thresholds combined with stacking to learn an optimal linear combination of C+T scores in a 10-fold cross-validation on the train set. The remaining variants are then pruned by discarding the ones with a P-value larger than a chosen significance level. Lassosum uses L1 and L2 regularization on the effect sizes and a linkage disequilibrium (LD) correlation matrix to penalize correlated and low-effect variants. The regularization coefficients were chosen by measuring model performance on the validation set. LDpred2 is a Bayesian method that uses a prior on effect sizes and an LD correlation matrix to re-weight effect estimates. LDpred2-auto (49) is a variant of LDpred2 in which two key model parameters, the SNP heritability and polygenicity, are estimated from the data. We used a shrinkage coefficient of 0.95 and 30 initial polygenicity parameters log-interpolated between 0.01 and 0.5. The LD correlation matrix was obtained from a reference panel (47). We used the validation set to identify optimal hyper-parameters such as P-value cutoffs and regularization coefficients for each method. We used the C+T, Lassosum2, and LDpred2 implementations of the bigsnpr (36) R package (version 1.9.10), using the default hyperparameters ranges for each PGS method.

### Network Architecture

We designed a deep learning neural network (DNN) to predict phenotypes from genetic data, illustrated in Figure 7. During training, SNPs are loaded in memory as a matrix of size *B* × *S*, where *B* represents the batch size and *S* is the number of SNPs. To reduce the dimensionality of the data, SNPs were filtered by a tunable p-value threshold *T*. The neural network’s architecture is an 8-layer pre-norm transformer (50) with two attention heads and GELU activation function (51). Similarly to vision transformers (50), we batched SNPs into arbitrary patches of length *L* to form a sequence of embeddings, using zero-padding to complete the last embedding. Inputs were then linearly mapped to match the input size of the transformer (512 in all experiments), and a vector containing covariate information was added as the first embedding of the sequence.

**Figure 7.**
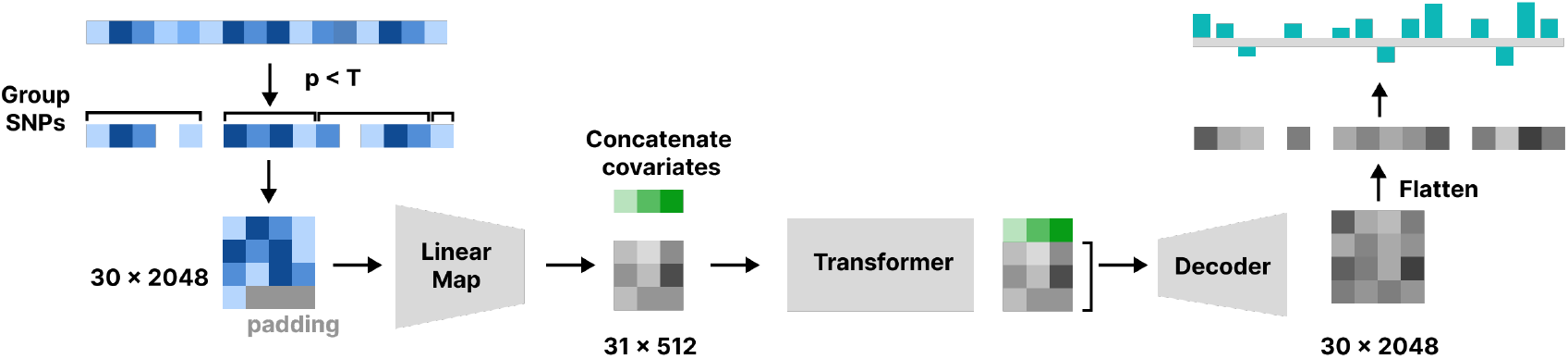
Overview of the architecture of the neural network. HM3 SNPs are first filtered by P-value threshold from the GWAS summary statistics. Then, SNPs are combined into blocks of size 2048 and linearly mapped to form embeddings for the transformer. Outputs from the transformer are then linearly decoded to the original input dimensions. The P-value threshold dictates the number of input blocks in the transformer, which is between 10 and 30, depending on the phenotype.

We used the effect sizes from LDpred2 to train the neural network in all our experiments. We used a batch size of *B* = 512, transformer input size of 512, feed-forward dimension of 512, and 0.3 dropout during training. The covariate vector included the same covariates from the GWAS for each trait. The patch size (*L* ∈ {128, 2048}), P-value thresholds (range 0.01-10^*−*6^), and learning rate (range 0.05-5 · 10^*−*4^) were individually tuned for each trait and were the only parameters that varied between traits. For a specific trait, we used the same patch size and p-value threshold for each data split. We used a linear decay for the learning rate with 300 warmup steps. Models were trained on a single NVIDIA GeForce RTX 3090 (24 GB) and were composed of approximately 14M parameters. We used the AdamW optimizer (52) with *ϵ* = 10 · 10^*−*8^, *β* = (0.9, 0.999). Training averaged between 8 to 12 hours for each trait. Inference time on the test set took approximately half an hour per phenotype.

For all phenotypes, we used explained variance (Equation 4) as the evaluation metric:

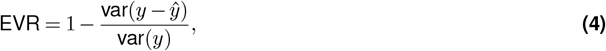

where *y* is the ground truth and *ŷ* is the prediction. We used this metric to select the best-performing model on the validation set and for the final evaluation of the held-out test set. We used smooth L1 loss (53) during training.

Interestingly, we observed different convergence patterns for each phenotype. Some, like BMI and SBP, tended to converge after one epoch despite a very low learning rate and even overfit after this point. On the other hand, height required a much larger learning rate and converged after around 20 epochs. We tuned the patch size such that the sequence length lay between 20 and 70, keeping patch sizes multiples of 2 between 256 and 2048. Hyperparameters that differed between phenotypes, namely the learning rate and the P-value threshold, are available in the code. Hyperparameters were kept the same for all data splits.

### Covariate Model

As some covariates can greatly impact the phenotype (for example, sex explains the majority of the variance for height), we found that directly using the phenotype as a ground truth would make the neural network diverge during training for some phenotypes. To solve this problem, we used another model that only used the covariates as input to predict the phenotype and trained the deep neural network on the residuals. We chose XGBoost(33), a gradient-boosted trees method that is the basis for method we used to benchmark (18), without the addition of high-impact SNPs. To be consistent, we used the same hyperparameters across data splits and phenotypes with 3-fold cross-validation for optimal model selection. For the XGBoost hyperparameters, we used a maximum depth of 5, *α* = 0, *γ* = 0, *η* = 0.01, a subsample of 80%, and a minimum chid weight of 10 in all our experiments. The weighted sum of effect estimates with P-values lower than 0.05 but higher than the P-value threshold of the deep neural network was then added back to the output of the covariate model. Finally, The DNN predictions *y*_*DNN*_ were linearly combined with the covariate model to form the final prediction.

### Data Loading During Training

Gene sequence variations formats such as bgen and pgen are compressed and optimized to query a single variant at a time to enable fast GWAS analysis. For our purposes, we needed a format that could allow us to efficiently load in memory all HM3 variants for a small number of subjects. We chose to convert the HM3 SNPs in bgen format to a Hierarchical Data Format (HDF5) with a Python script using the h5py (54) and bgen_reader (55) libraries. When loading the data, we implemented an efficient dataloader that merges genotype and phenotype information. This data format allowed us to load a batch of 512 samples containing 1.1M SNPs in memory in less than 10 ms, which was acceptable for training. Bgens were converted to a single HDF file with a Python script, which only needed to be done once. The code for the dataloader and bgen conversion is available online (see section Code Availability) and can be reused for other projects. We encoded allele counts as 0, 1, and 2 for homozygous reference, heterozygous alternative and homozygous alternative allele. The samples were ordered as in the sample file from the.bgen of the first chromosome.

### Adding in low effects as constants

To keep the inputs’ dimensionality relatively low and avoid including extremely small effects, we summed up the effects that were lower than 0.05% of the maximum and only included the others in the input of the neural network. The sum of the smaller effects was then added to the *y*_*DNN*_ output. Assuming that the deep neural network output only zeros, the network’s architecture is such that output would be the same as the LDpred2 weighted sum. Additionally, we experimented with adding these residual PGS as an additional covariate, which we found improved results on predictions from REGENIE effect estimates.

### Model performance evaluation and comparison to existing methods

We compared the performance of our approach to three other benchmark methods. We used the polygenic risk score predictions from LDpred2 as input for each method and included the same covariates as our approach to ensure fair comparison. It was recently found (18) that including high-impact SNPs in a non-linear model can increase the quality of genetic prediction. We modified this existing method to be computationally feasible while enabling fair comparison. To be precise, instead of filtering SNPs with LASSO regression before XGboost, which we found to be computationally expensive due to the size of UKB, we filtered them by P-value thresholding. We considered for inclusion in the model all SNPs with a *p* value *<* 10^*−*4^ using our GWAS summary statistics for the corresponding trait, keeping the same data splits as previously described. We then used eight relative thresholds *α* values between 0 and 1 and kept SNPs with a P-value in the top *T* percentile as additional inputs. We verified that adding additional SNPs as input would not result in any further prediction performance gain, see Supplementary section 6.

We fitted XGBoost and LASSO models by including covariates (sex, age, first 20 PCs), selected SNPs, and the LDpred2 risk score prediction. We selected the model that minimized the MSE for each phenotype. For XGBoost, we always used a learning rate of 0.01, maximum depth of 5, minimum child weight of 10, and subsample of 80%. Each model was fit using 3-fold cross-validation on the training set, allowing up to 2000 boosted trees with early stopping after 20 rounds. We repeated this process for all 10 traits and 3 data splits. Analysis was conducted using Python 3 and the scikit-learn (56) and xgboost (33) packages.

## Data Availability

Data referred to in the present work is available by application at https://www.ukbiobank.ac.uk/.

## Data Analysis with R

Data analysis was performed with publicly available packages: tidyverse v1.3.1 (57), and dplyr v1.0.8 (58).

## Data Analysis with Python

The covariate model was implemented using the xgboost python library (33). The deep learning model was implemented in Pytorch (59) version 2.3.

## Code Availability

Code used for processing genetic data, GWAS analyses, and training of the neural network for this manuscript is provided on a dedicated GitLab repository https://gitlab.com/cgeo/delphi.

## Data availability

No data were generated in the present study. UK Biobank data are publicly available by application (https://www.ukbiobank.ac.uk/enable-your-research/register).

## Acknowledgements

This research has been conducted using the UK Biobank resource under application number 80108, with funding from the Swiss National Science Foundation (Sinergia CRSII5_202276/1).

