## Supplementary material for "Delphi: Deep Learning for Polygenic Risk Prediction"

### Supplementary Note 1: Baseline PGS results

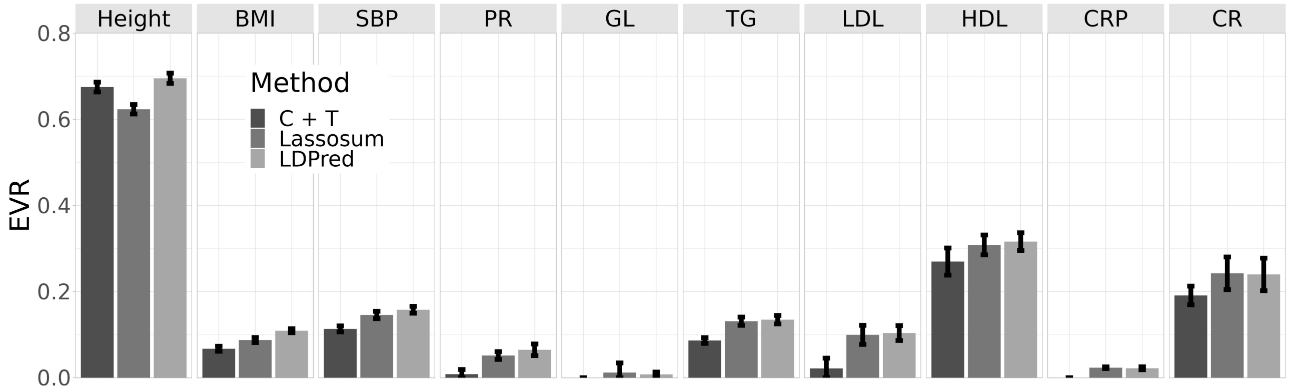


**Figure 1.** C+T, Lassosum2 and LDpred2 linear PGS results on the validation set. We show the best-performing model of three independent data splits. Validation sets were used to determine the optimal parameters for each method. Error bars indicate the standard deviation between splits. EVR: explained variance, BMI: Body mass index, CRP: C-Reactive protein, GL: glucose, LDL: low-density lipoproteins, SBP: systolic blood pressure.

### Supplementary Note 2: Other Performance Metrics

We also show the performance of Delphi for continuous phenotypes in terms of mean absolute error 2. Delphi also consistently outperforms other methods w.r.t this metric.

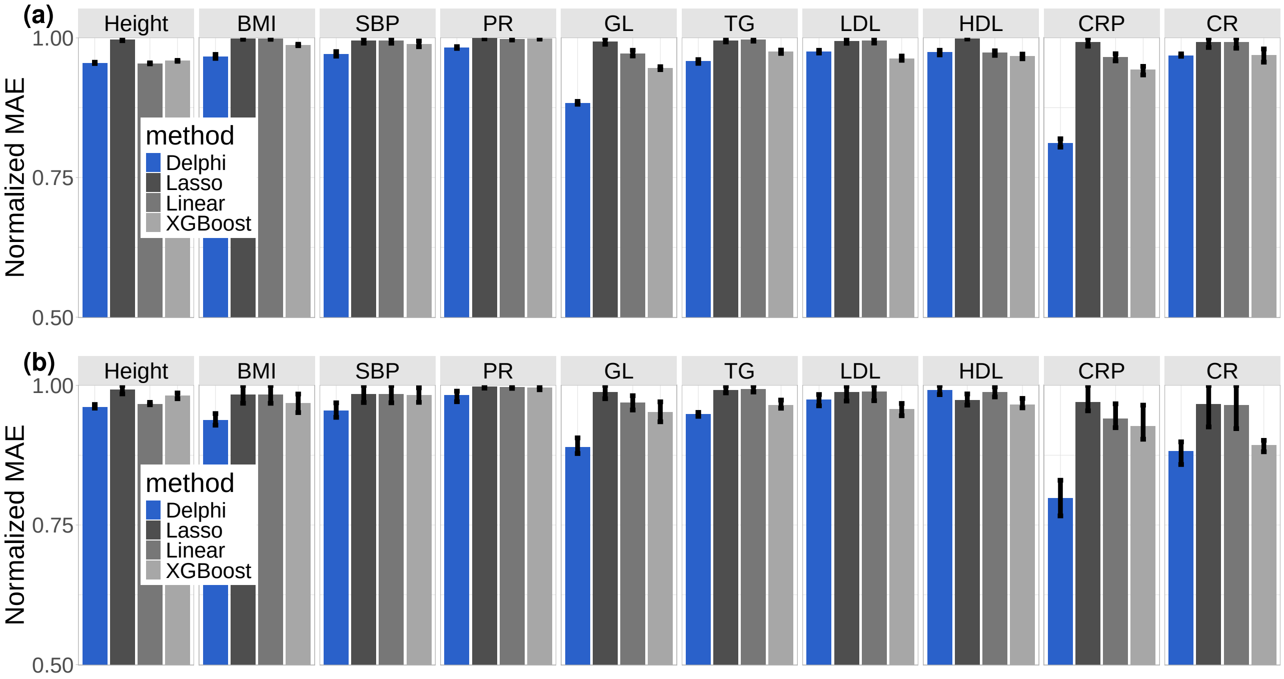


**Figure 2.** Normalized mean absolute error (divided by the maximum observed MAE across experiments - lower is better) of polygenic predictions for five phenotypes in the UK Biobank. a) All participants; b) Individuals from non-British white ancestry. We report results for a linear PGS model, lasso regression, an XGBoost model including the dosage of multiple high-impact SNPs as input, and our method. See Figure 4 for acronyms.

### Supplementary Note 3: Additional Performance Comparison

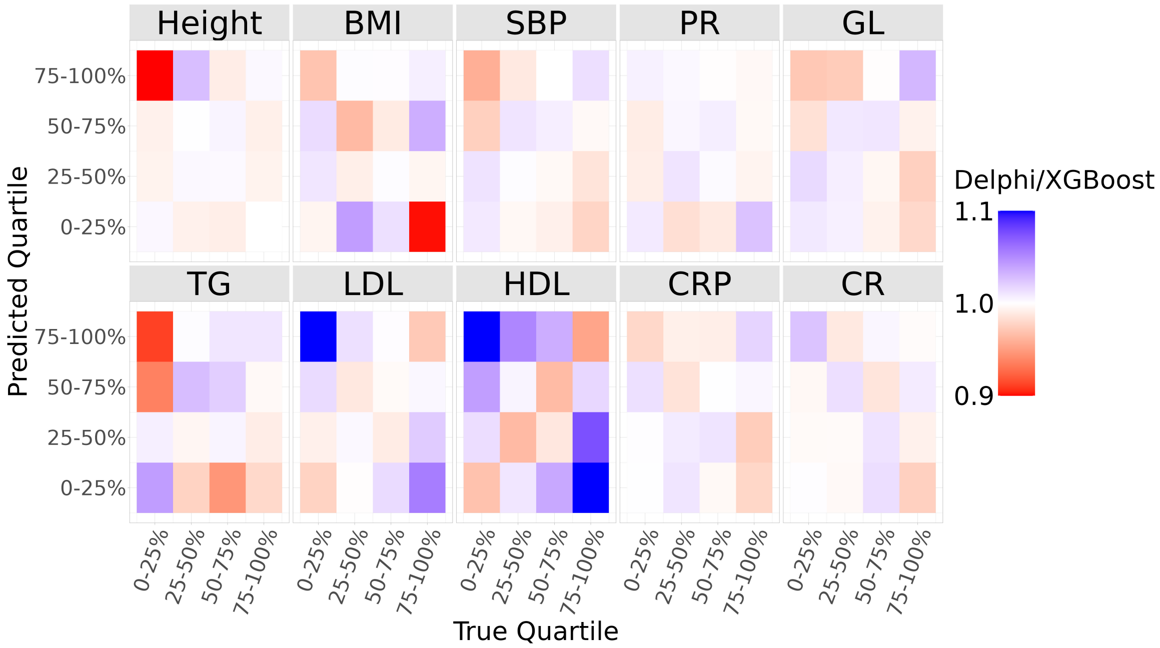


**Figure 3.** Ratio of quartile distributions of predictions between Delphi and XGBoost, on the test set for five phenotypes. Although the proportion of correctly binned subjects (same predicted and ground-truth quartiles) is similar for both methods, Delphi tends to avoid extreme differences between prediction and ground-truth. Values for height were bounded between 0.9 and 1.1 for visibility; original values are in the range of 0.2 and 1.3.

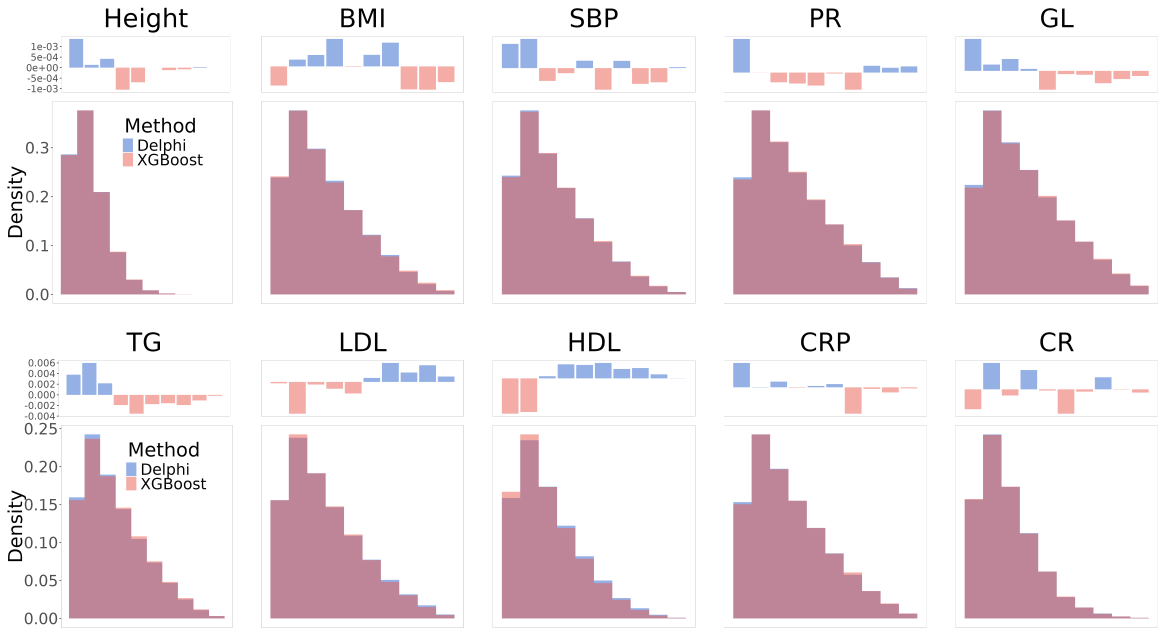


**Figure 4.** Histogram of the absolute difference between predicted and true deciles on the test set for five phenotypes. Delphi consistently bins subjects more adequately than XGBoost for most phenotypes.

### Supplementary Note 4: Performance on other Ancestries

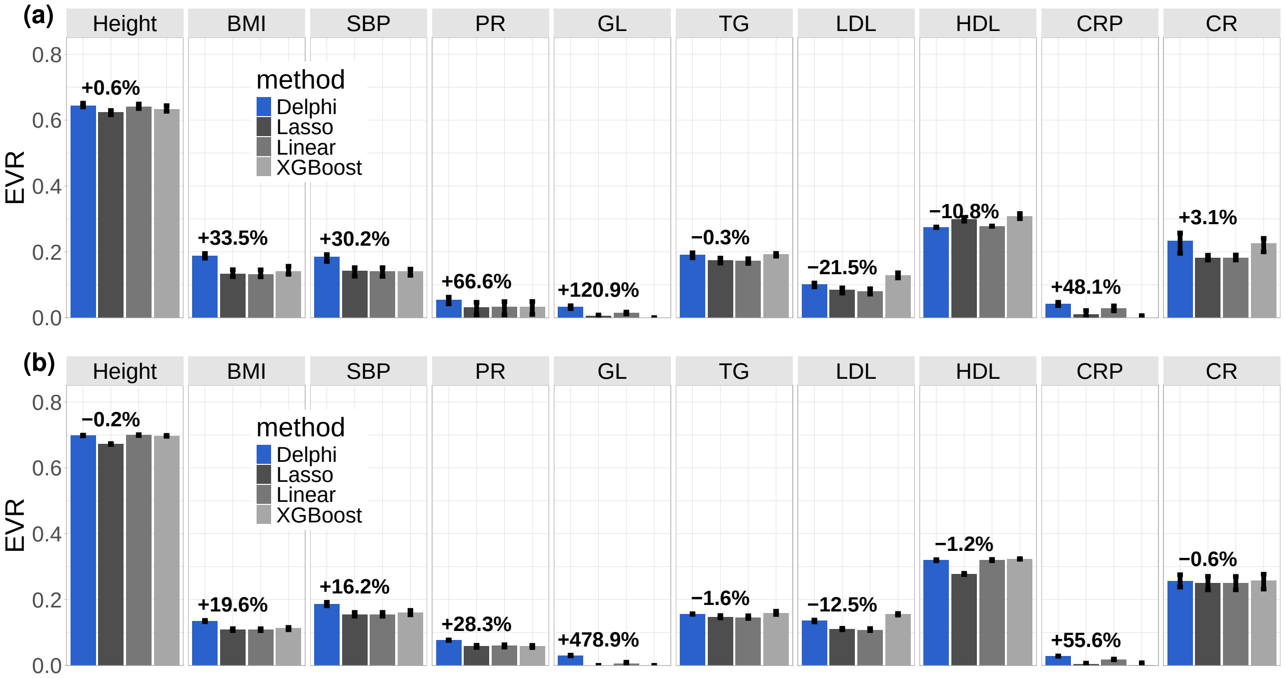


**Figure 5.** Accuracy of polygenic predictions for ten phenotypes in the UK Biobank depending on ancestry. a) predictions for ten phenotypes in the UK Biobank on individuals with non-British white ancestry. b) Prediction results for individuals with British white ancestry. We report results for a linear PGS model, lasso regression, and XGBoost, including the allele count of multiple high-impact SNPs as input, and our method.

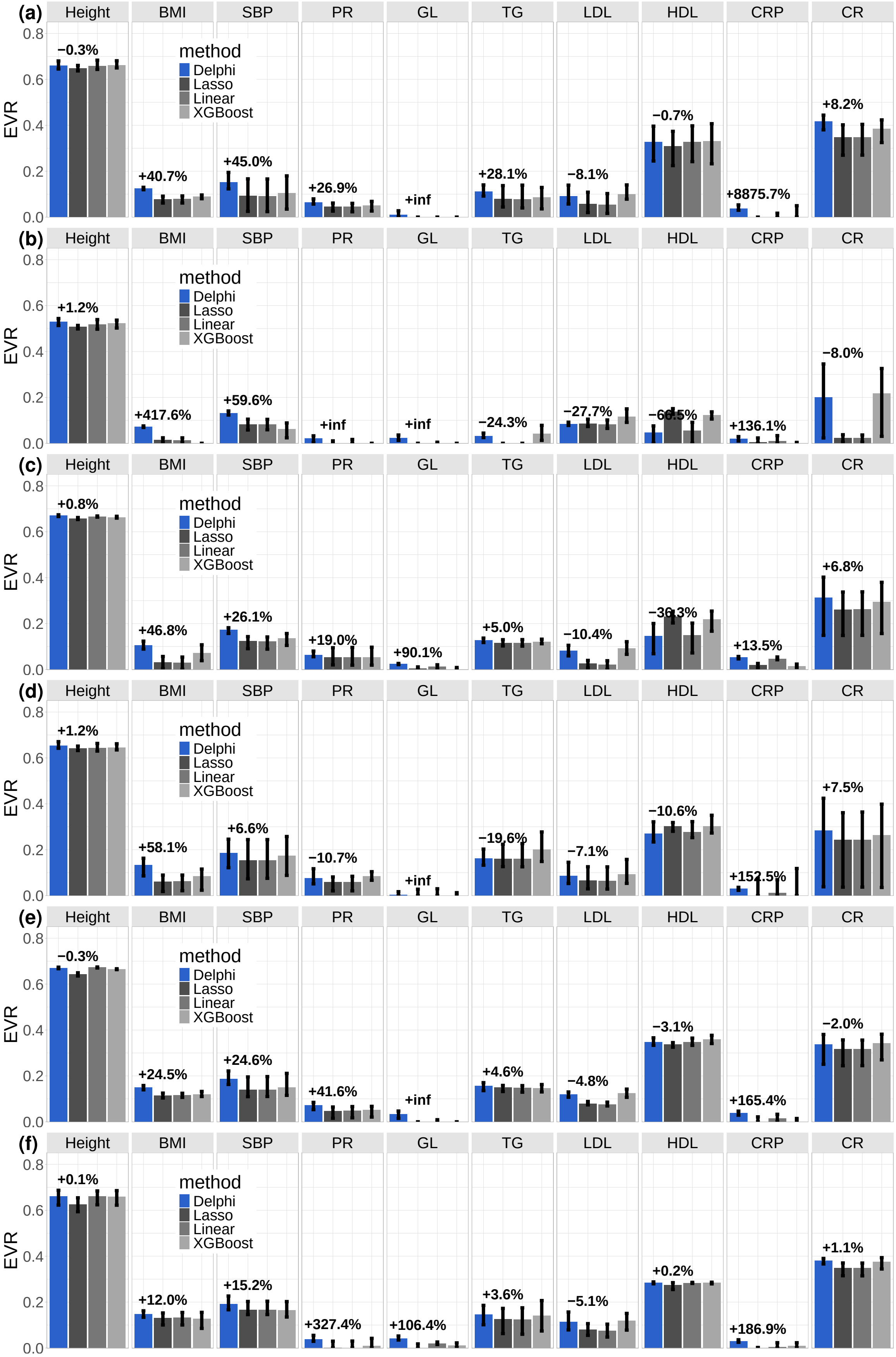


**Figure 6.** Accuracy of polygenic predictions for ten continuous phenotypes in the UK Biobank for six self-reported ancestries. From top to bottom: a) Askenazi, b) Caribbean, c) India, d) Iran, d) Italy, e) Poland.

### Supplementary Note 5: Prediction Performance Tables

Table of prediction performance for results in Figure 3 and Figure 5 in the main text.

| | Phenotype | Delphi | Lasso | Linear | XGBoost |
|---|---|---|---|---|---|
| | BMI | 0.14 (0.14–0.14) | 0.11 (0.11–0.11) | 0.11 (0.11–0.11) | 0.11 (0.11–0.12) |
| | Height | 0.70 (0.70–0.70) | 0.67 (0.67–0.67) | 0.70 (0.70–0.70) | 0.69 (0.69–0.69) |
| | SBP | 0.19 (0.18–0.19) | 0.16 (0.15–0.16) | 0.16 (0.15–0.16) | 0.16 (0.15–0.17) |
| | LDL | 0.13 (0.13–0.14) | 0.11 (0.11–0.11) | 0.11 (0.10–0.11) | 0.15 (0.15–0.16) |
| All | GL | 0.03 (0.03–0.03) | 0.00 (0.00–0.00) | 0.01 (0.00–0.01) | 0.00 (0.00–0.00) |
| | CRP | 0.03 (0.03–0.03) | 0.00 (0.00–0.01) | 0.02 (0.02–0.02) | 0.00 (0.00–0.01) |
| | HDL | 0.32 (0.32–0.32) | 0.28 (0.28–0.28) | 0.32 (0.32–0.32) | 0.32 (0.32–0.32) |
| | PR | 0.07 (0.07–0.08) | 0.06 (0.05–0.06) | 0.06 (0.05–0.06) | 0.06 (0.05–0.06) |
| | CR | 0.25 (0.24–0.27) | 0.24 (0.22–0.26) | 0.24 (0.22–0.26) | 0.25 (0.23–0.27) |
| | TG | 0.16 (0.16–0.16) | 0.15 (0.15–0.15) | 0.15 (0.14–0.15) | 0.16 (0.16–0.17) |

| | Phenotype | Delphi | Lasso | Linear | XGBoost |
|---|---|---|---|---|---|
| | BMI | 0.10 (0.08–0.10) | 0.00 (0.00–0.01) | 0.00 (0.00–0.00) | 0.00 (0.00–0.02) |
| | Height | 0.50 (0.45–0.52) | 0.49 (0.45–0.51) | 0.48 (0.44–0.51) | 0.49 (0.45–0.52) |
| | SBP | 0.11 (0.08–0.14) | 0.07 (0.03–0.13) | 0.07 (0.03–0.13) | 0.03 (0.00–0.10) |
| | LDL | 0.02 (0.01–0.03) | 0.01 (0.00–0.02) | 0.01 (0.00–0.02) | 0.06 (0.06–0.07) |
| Nigeria | GL | 0.02 (0.02–0.02) | 0.01 (0.00–0.04) | 0.02 (0.00–0.04) | 0.00 (0.00–0.03) |
| | CRP | 0.03 (0.02–0.04) | 0.00 (0.00–0.00) | 0.01 (0.00–0.01) | 0.00 (0.00–0.00) |
| | HDL | 0.01 (0.00–0.03) | 0.13 (0.11–0.16) | 0.02 (0.00–0.04) | 0.12 (0.11–0.15) |
| | PR | 0.00 (0.00–0.02) | 0.00 (0.00–0.00) | 0.00 (0.00–0.00) | 0.00 (0.00–0.00) |
| | CR | 0.24 (0.07–0.35) | 0.08 (0.00–0.28) | 0.09 (0.00–0.29) | 0.21 (0.05–0.31) |
| | TG | 0.00 (0.00–0.01) | 0.00 (0.00–0.00) | 0.00 (0.00–0.00) | 0.00 (0.00–0.03) |

| | Phenotype | Delphi | Lasso | Linear | XGBoost |
|---|---|---|---|---|---|
| | BMI | 0.11 (0.09–0.12) | 0.03 (0.00–0.06) | 0.03 (0.00–0.05) | 0.07 (0.04–0.11) |
| | Height | 0.67 (0.67–0.67) | 0.66 (0.65–0.66) | 0.67 (0.66–0.67) | 0.66 (0.66–0.67) |
| | SBP | 0.17 (0.16–0.18) | 0.12 (0.09–0.14) | 0.12 (0.09–0.14) | 0.14 (0.10–0.16) |
| | LDL | 0.08 (0.06–0.11) | 0.03 (0.01–0.04) | 0.02 (0.00–0.04) | 0.09 (0.07–0.12) |
| India | GL | 0.02 (0.02–0.03) | 0.00 (0.00–0.01) | 0.01 (0.00–0.02) | 0.00 (0.00–0.01) |
| | CRP | 0.05 (0.05–0.06) | 0.02 (0.01–0.03) | 0.05 (0.04–0.05) | 0.01 (0.01–0.02) |
| | HDL | 0.15 (0.07–0.20) | 0.23 (0.20–0.25) | 0.15 (0.07–0.20) | 0.22 (0.17–0.25) |
| | PR | 0.06 (0.06–0.08) | 0.05 (0.02–0.09) | 0.05 (0.02–0.10) | 0.05 (0.02–0.10) |
| | CR | 0.31 (0.15–0.40) | 0.26 (0.15–0.34) | 0.26 (0.15–0.34) | 0.29 (0.16–0.38) |
| | TG | 0.13 (0.12–0.14) | 0.12 (0.10–0.13) | 0.12 (0.10–0.13) | 0.12 (0.11–0.13) |

| | Phenotype | Delphi | Lasso | Linear | XGBoost |
|---|---|---|---|---|---|
| | BMI | 0.10 (0.06–0.13) | 0.00 (0.00–0.03) | 0.00 (0.00–0.03) | 0.10 (0.08–0.12) |
| | Height | 0.56 (0.52–0.60) | 0.54 (0.48–0.59) | 0.54 (0.51–0.59) | 0.55 (0.51–0.60) |
| | SBP | 0.24 (0.23–0.25) | 0.21 (0.20–0.22) | 0.21 (0.20–0.22) | 0.23 (0.23–0.25) |
| | LDL | 0.14 (0.12–0.16) | 0.10 (0.07–0.12) | 0.10 (0.07–0.12) | 0.15 (0.15–0.15) |
| China | GL | 0.03 (0.02–0.05) | 0.00 (0.00–0.00) | 0.00 (0.00–0.02) | 0.00 (0.00–0.00) |
| | CRP | 0.00 (0.00–0.03) | 0.00 (0.00–0.00) | 0.00 (0.00–0.00) | 0.00 (0.00–0.03) |
| | HDL | 0.28 (0.26–0.31) | 0.26 (0.23–0.29) | 0.29 (0.27–0.32) | 0.30 (0.27–0.32) |
| | PR | 0.06 (0.03–0.08) | 0.00 (0.00–0.03) | 0.00 (0.00–0.03) | 0.03 (0.00–0.07) |
| | CR | 0.44 (0.42–0.45) | 0.23 (0.06–0.40) | 0.23 (0.06–0.41) | 0.45 (0.42–0.48) |
| | TG | 0.12 (0.09–0.15) | 0.12 (0.07–0.15) | 0.11 (0.06–0.14) | 0.16 (0.10–0.22) |

### Supplementary Note 6: Validation on the number of input SNPs

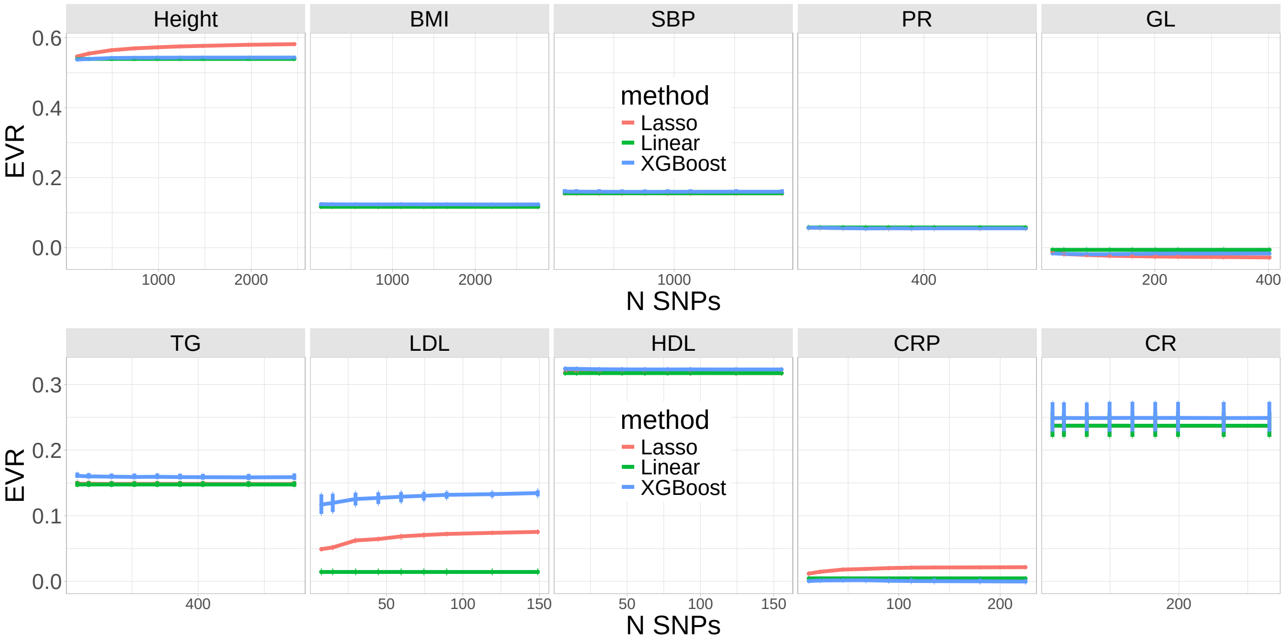


**Figure 7.** Performance of benchmarked methods w.r.t the number of high-impact SNPs added as input. Each method also included sex, age, the first 20 principal components, and a PGS prediction from LDpred2 as input.

### Supplementary Note 7: REGENIE Predictions

We extensively benchmarked our method using summary statistics from REGENIE. Benchmarked methods were the same as described in section Model performance evaluation and comparison to existing methods, and used the PGS predictions and high-impact SNPs from REGENIE. The same data splits and covariates were used. We observed an increase across the three data splits in the percentage of variance explained for height, BMI, glucose, C reactive protein, HDL, and creatinine, as shown on Figure 8. Our method performed similarly on pulse rate and systolic blood pressure and worse on triglycerides and LDL. This drop in performance in some phenotypes could vanish with more extensive parameter tuning. We have slightly the inputs as described in Methods section Adding in low effects as constants (line 370), as we saw that this provided slight performance improvements with REGENIE weights. We also observed a slight drop in predictive performance for individuals of non-British white ancestry, as shown on Figure 9. This may be due to the fact that REGENIE weights serve as a better baseline for individuals in the majority group and could be fixed by experimenting with a wider range of P-value thresholds. Additionally, multiple sources of effect estimates could be included concurrently, although we leave those experiments for further research.

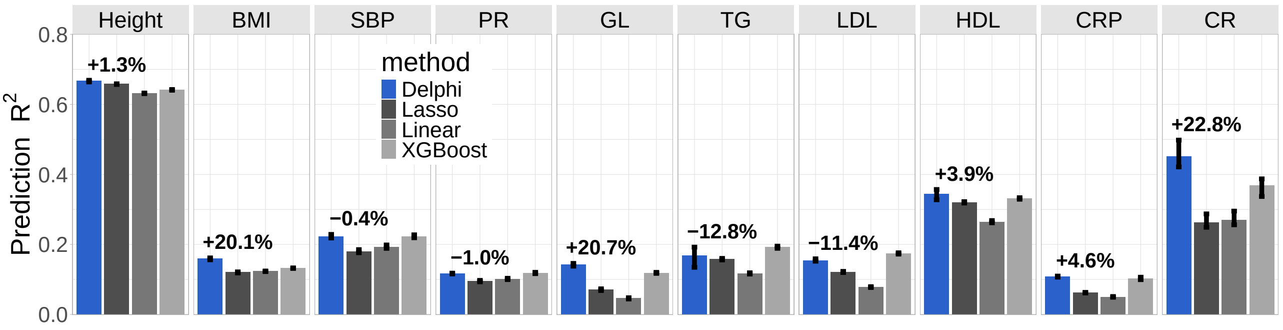


**Figure 8.** Accuracy of polygenic predictions using summary statistics from REGENIE for ten phenotypes in the UK Biobank. We report results for a linear PGS model, lasso regression, and XGBoost models, including the allele count of multiple high-impact SNPs as input, and our method.

| Phenotype | Delphi | Lasso | Linear | XGBoost |
|---|---|---|---|---|
| BMI | 0.16 (0.16–0.16) | 0.12 (0.12–0.12) | 0.12 (0.12–0.12) | 0.13 (0.13–0.13) |
| Height | 0.67 (0.66–0.67) | 0.66 (0.66–0.66) | 0.63 (0.63–0.63) | 0.64 (0.64–0.64) |
| SBP | 0.22 (0.22–0.23) | 0.18 (0.18–0.19) | 0.19 (0.19–0.20) | 0.22 (0.22–0.23) |
| LDL | 0.15 (0.15–0.16) | 0.12 (0.12–0.12) | 0.08 (0.08–0.08) | 0.17 (0.17–0.18) |
| GL | 0.14 (0.14–0.15) | 0.07 (0.07–0.07) | 0.05 (0.04–0.05) | 0.12 (0.12–0.12) |
| CRP | 0.11 (0.11–0.11) | 0.06 (0.06–0.06) | 0.05 (0.05–0.05) | 0.10 (0.10–0.11) |
| HDL | 0.34 (0.33–0.36) | 0.32 (0.32–0.32) | 0.26 (0.26–0.27) | 0.33 (0.33–0.33) |
| PR | 0.12 (0.12–0.12) | 0.09 (0.09–0.10) | 0.10 (0.10–0.10) | 0.12 (0.12–0.12) |
| CR | 0.45 (0.42–0.50) | 0.26 (0.25–0.29) | 0.27 (0.26–0.30) | 0.37 (0.34–0.39) |
| TG | 0.17 (0.13–0.19) | 0.16 (0.16–0.16) | 0.12 (0.12–0.12) | 0.19 (0.19–0.20) |

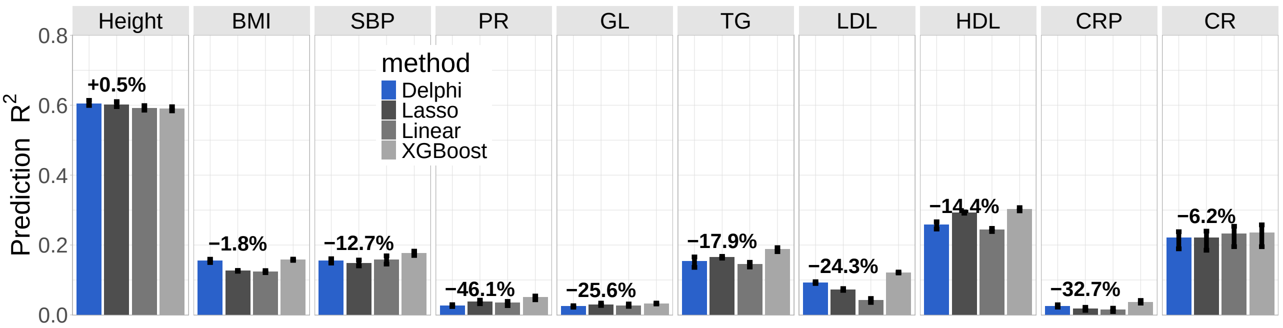


**Figure 9.** Accuracy of polygenic predictions using summary statistics from REGENIE for ten phenotypes in the UK Biobank on individuals from non-British white ancestry. We report results for a linear PGS model, lasso regression, and XGBoost models, including the allele count of multiple high-impact SNPs as input, and our method.

### Supplementary Note 8: Additional Synthetic Experiments

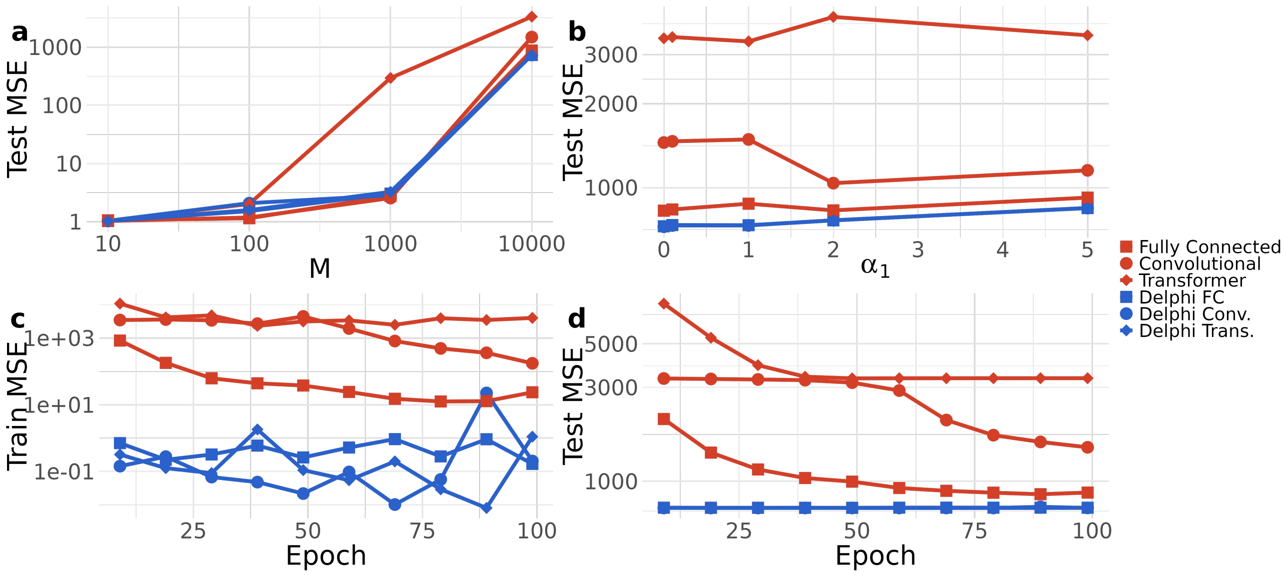


**Figure 10.** Performance comparison between methods (direct prediction and Delphi) for three different network architectures. a) Mean squared error (MSE) on the test set divided by the number of features w.r.t. the number of features ($\alpha_1$ = 1). b) Test set MSE w.r.t the interaction factor for $M = 10'000$ features. c) MSE on the training set during training for $M = 10'000$ features and d) MSE on the test set. Delphi achieves a lower test MSE for the simulated additive phenotype and converges much faster (approximatively 10 v.s. 100+ epochs) when the number of features is large.

We conducted synthetic experiments with a phenotype that was not polygenic to showcase the limitations of our method. The phenotype is constructed with the following equation:

$$\begin{aligned} \mathbf{y} &= \mathbf{X_0} + \mathbf{X_0} \odot \mathbf{X_0}\beta + \epsilon \\ \beta &\sim \mathcal{N}(\mathbf{0}, \alpha_1), \\ \epsilon &\sim \mathcal{N}(\mathbf{0}, \alpha_2) \end{aligned} \tag{5}$$

where $\mathbf{y} \in \mathbb{R}^N$ is the synthetic phenotype for $N$ subjects, $\mathbf{X} \in \{0,1,2\}^{N \times M}$ is a matrix of randomly drawn integers between 0 and 2, with $N$ subjects and $M$ variants. $\mathbf{X_0}$ represents the first variant, the only one that has an effect on the phenotype. We chose $\alpha_1 = 1$ and $\alpha_2 = 0.1$. As shown in Figure 12, in this case, Delphi does not produce any meaningful predictions despite having a lower training set MSE. Thus, this method is only applicable with phenotypes that are highly polygenic.

#### A. HAPNEST Experiments

We provide additional details on the construction of synthetic genotypes and phenotypes using HAPNEST (34), as well as simulation parameters not described in the main text. Genotypes were generated using HAPNEST, which simulates individual-level genetic data by resampling haplotype segments from a reference panel under a Li and Stephens–style model of linkage disequilibrium. Synthetic haplotypes are constructed as mosaics of reference haplotypes, with segment lengths governed by population-specific recombination rates and effective population sizes. Coalescent times are sampled for each segment and combined with mutation age estimates to determine whether variants are copied, thereby limiting overfitting to the reference panel while preserving realistic LD structure.
Phenotypes were generated using the HAPNEST phenotype module, in which genetic effects are modeled as weighted sums of causal allele counts. For each causal SNP $i$, the effect size $b_i$ is drawn from a zero-mean Gaussian distribution whose variance depends on minor allele frequency, local linkage disequilibrium, and functional annotations.

$$b_i \sim \mathcal{N}\Big(0,\ [p_i(1-p_i)]^a\, r_i^b s_i^c\Big) \tag{6}$$

where $p_i$ denotes the minor allele frequency of SNP $i$, $r_i$ aptures local linkage structure, and $s_i$ denotes functional annotation weights. The exponents $a, b$ and $c$ control the strength of each contribution and were set to the HAPNEST default values ($a = -0.4, b = -1, c = 0.5$). Polygenicity controls the fraction of SNPs assigned non-zero effects, which we varied in our experiments. The proportion of covariate influence was set to $0.2$, and correlation between phenotypes to 0. All other parameters were set to the default values.

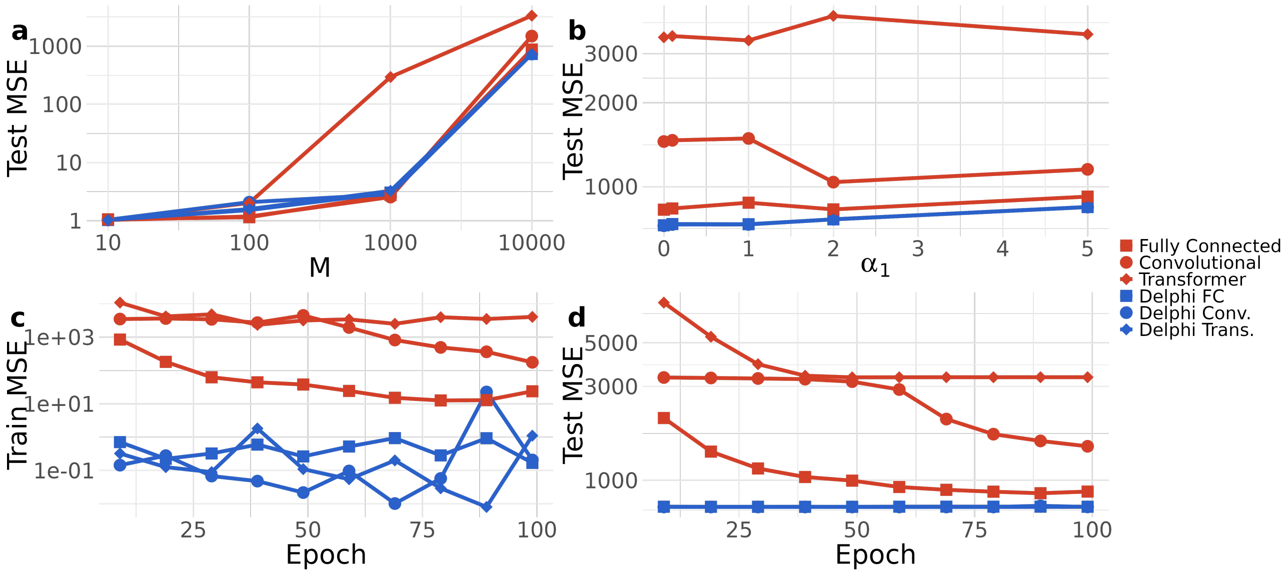


**Figure 11.** Performance comparison between methods (direct prediction and Delphi) for three different network architectures. Phenotypes were generated based on Equation 5. Mean squared error (MSE) on the test set divided by the number of features w.r.t. a) the number of features ($\alpha_1$ = 1) b) the interaction factor $\alpha_1$. We also report the loss per training epoch on c) the train set and d) the test set. The dataset is of size $N = 10'000$. Delphi achieves a lower test MSE for the simulated additive phenotype and converges much faster (approximatively 10 v.s. 100+ epochs)

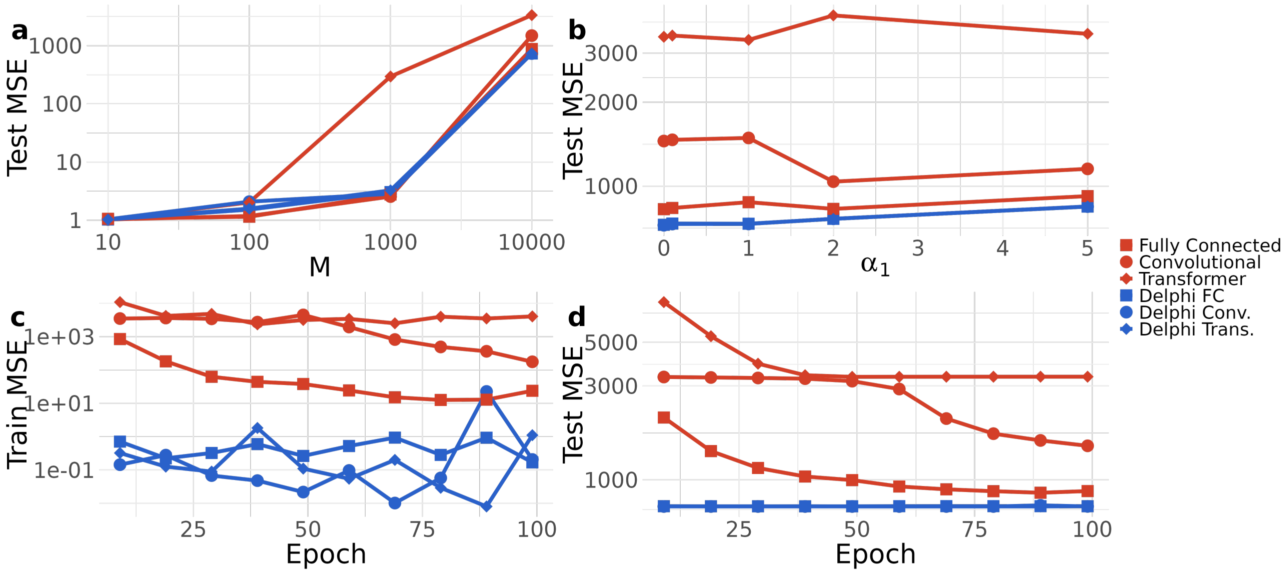


**Figure 12.** Train and test set mean squared error (MSE) during training for Delphi and benchmark methods for a non-polygenic synthetic phenotype. Delphi does not output meaningful prediction in this setup.
